# Ensuring a successful transition from Pap to HPV-based primary screening in Canada: a study protocol to investigate the psychosocial correlates of women’s screening intentions

**DOI:** 10.1101/2022.03.28.22272903

**Authors:** Gabrielle Griffin-Mathieu, Ben Haward, Ovidiu Tatar, Patricia Zhu, Samara Perez, Gilla K. Shapiro, Emily McBride, Erika L. Thompson, Laurie W. Smith, Aisha K. Lofters, Ellen M. Daley, Juliet R. Guichon, Jo Waller, Marc Steben, Kathleen M. Decker, Marie-Hélène Mayrand, Julia M. L. Brotherton, Gina S. Ogilvie, Gregory D. Zimet, Teresa Norris, Zeev Rosberger

**Affiliations:** Lady Davis Institute for Medical Research (LDI), Jewish General Hospital, Montreal, QC, Canada; Research Center, Centre Hospitalier de l’Université de Montréal (CRCHUM), Montreal, QC, Canada; Department of Psychiatry, McGill University, Montreal, QC, Canada; McGill University Health Centre (MUHC), Montreal, QC, Canada; Department of Oncology, McGill University, Montreal, QC, Canada; Department of Supportive Care, Princess Margaret Cancer Centre, University Health Network, Toronto, ON, Canada; Department of Behavioural Science & Health, University College London, London, United Kingdom; Department of Biostatistics and Epidemiology, School of Public Health, The University of North Texas Health Science Center, Fort Worth, TX, USA; BC Cancer Agency, Vancouver, BC, Canada; Department of Family and Community Medicine, University of Toronto, Toronto, ON, Canada; Department of Family and Community Medicine, Women’s College Hospital, Toronto, ON, Canada; College of Public Health, University of South Florida, Tampa, FL, USA; Departments of Community Health Sciences and Pediatrics, University of Calgary, Calgary, AB, Canada; School of Cancer and Pharmaceutical Sciences, King’s College London, London, United Kingdom; School of Public Health, l’Université de Montréal, Montreal, QC, Canada; Department of Community Health Sciences, University of Manitoba, Winnipeg, MB, Canada; CancerCare Manitoba Research Institute, Winnipeg, MB, Canada; Département d’obstétrique-gynécologie, Université de Montréal, Montreal, QC, Canada; Melbourne School of Population and Global Health, University of Melbourne, Melbourne, Victoria, Australia; Population Health, Australian Centre for the Prevention of Cervical Cancer, Melbourne, Victoria, Australia; Faculty of Medicine, University of British Columbia, Vancouver, BC, Canada; BC Women’s Hospital, Vancouver, BC, Canada; School of Medicine, Indiana University, IN, USA; HPV Global Action, Montreal, QC, Canada; Department of Psychology, McGill University, Montreal, QC, Canada

## Abstract

**Introduction:** The Human Papillomavirus (HPV) test has emerged as a significant improvement over cytology for primary cervical cancer screening. In Canada, provinces and territories are moving towards implementing HPV testing in cervical cancer screening programs. While an abundance of research exists on the benefits of HPV-based screening, there is a dearth of research examining women’s understanding of HPV testing. In other countries, failure to adequately address women’s concerns about changes has disrupted implementation of HPV-based screening. This study protocol describes a multi-step approach to develop psychometrically valid measures and to investigate psychosocial correlates of women’s intentions to participate in HPV-based cervical cancer screening.

**Materials and Methods:** We conducted a web-based survey of Canadian women to assess the acceptability and feasibility of a questionnaire, including validation of scales examining: cervical cancer knowledge, HPV testing knowledge, HPV testing attitudes and beliefs, and HPV test self-sampling attitudes and beliefs. Preferences for cervical cancer screening were assessed using Best-Worst Scaling methodology.

A second web-based survey will be administered to a national sample of Canadian women in June-July of 2022 using the validated scales. Differences in the knowledge, attitudes, beliefs, and preferences of women who are currently either underscreened or adequately screened for cervical cancer will be examined through bivariate analyses. Multinomial logistic regression will be used to estimate the associations between psychosocial and sociodemographic factors and intentions to screen using HPV-based screening.

**Study Impact and Dissemination:** Findings will provide direction for Canadian public health authorities to align guidelines to address women’s concerns and optimize acceptability and uptake of HPV-based primary screening. Validated scales can be used by other researchers to improve and standardize measurement of psychosocial factors impacting HPV test acceptability. Study results will be disseminated through peer-reviewed journal articles, conference presentations, and direct communication with researchers, clinicians, policymakers, media, and specialty organizations.

## Introduction

Cervical cancer is the fourth most common cancer in women and presents a significant risk to all people with a cervix [1-3]. In 2018, an estimated 570,000 women were newly diagnosed with cervical cancer worldwide, with 311,000 deaths from the disease [4]. In Canada, more than 1,300 women are diagnosed with, and over 400 die from cervical cancer each year [5]. Cytology testing using the Papanicolaou smear, commonly referred to as the Pap test, allows for the detection and subsequent treatment of precancerous lesions that might lead to cervical cancer. Canadian women have been widely screened for cervical cancer using the Pap test for over fifty years [6]. There are guidelines in each province/territory that currently recommend screening every 1 to 3 years starting at age 21 or 25 [7], and most Canadian jurisdictions have organized screening programs [8].

Cervical cancer is caused by persistent infection with high risk Human Papillomavirus (HPV) types [9]. The HPV test, which detects high risk HPV DNA in cervical cells, has emerged as a significant improvement over cytology for cervical cancer screening. Compared to cytology-based screening, HPV-based screening has been shown to offer a 60-70% higher protection against the development of cervical cancer [10, 11]. The HPV test shows improved sensitivity and a negative test result has high negative predictive value, thus providing greater reassurance against cervical lesion development [12]. Furthermore, this allows for increased intervals between cervical screenings and reduced testing costs [12]. HPV testing also introduces the possibility for vaginal self-sampling by the patient. This is not possible with cytology, which requires cervical cell collection by a healthcare provider, a process that can be uncomfortable or invasive [13, 14]. Meta-analyses have shown that self-sampling has similar test accuracy when compared to healthcare provider administered sampling [15], and could increase uptake among those who are underscreened when provided as a screening option [15, 16].

For these reasons, HPV testing is now considered the preferred method of screening by the World Health Organization [17] and is recommended by multiple specialty organizations worldwide (e.g., United States Preventive Services Task Force [USPSTF] [18], European Society of Gynaecologic Oncology [ESGO], European Federation of Colposcopy [EFC] [19]). Several countries have implemented HPV-based organized screening programs (e.g., Australia, U.K., Netherlands), including the use of self-sampling as a collection option [20, 21]. However, program implementations have encountered challenges. For example, in Australia prior to the introduction of HPV testing, an online petition against the proposed changes gained widespread support. In an analysis of comments on the petition, Australian women felt that the policy “threatened or de-valued” women’s health, represented a government cost-cutting measure, and that increased screening intervals and later age of screening initiation would prevent early detection of cervical abnormalities [22]. Similarly, in Wales an increase in screening intervals associated with the shift to HPV-based screening has been met with public backlash, with over 1.2 million signatures on an online petition against the change at the time of writing [23]. While these guidelines are grounded in evidence, a disconnect has been observed between women’s and public health’s views of cervical cancer screening changes. These health authorities failed to effectively and proactively communicate why changes were warranted, why screening intervals might change, and what HPV test results indicate [22-26].

### Screening Landscape in Canada

In Canada, provinces and territories are in different planning phases for HPV-based cervical cancer screening programs [7, 27], and the nationwide introduction of HPV-based primary screening is a key priority of the recent Canadian Partnership Against Cancer (CPAC) Action Plan for the Elimination of Cervical Cancer [5]. While cytology-based screening is well-established in Canada, screening coverage has failed to reach the CPAC target of including ≥80% of women [28]. This suggests that innovative approaches are needed to reach those women who are *underscreened* (i.e., longer than three years since previous Pap test, or never screened). About 40% of women in Canada diagnosed with cervical cancer report either never having had a Pap test, or not having had one in more than three years [29]. Underscreened women are often members of ethnic, linguistic, gender, and sexual minorities or have lower socioeconomic status [30-33]. Only 65% of recent female immigrants report having a Pap test in the previous three years, and 26% of non-English or French speakers report never having a Pap test [29]. In a study of cancer screening registries in Ontario, transgender patients were significantly less likely than cisgender patients to be screened for cervical cancer (56% vs. 72%) [32], reflective of barriers faced by transgender and other gender diverse people in seeking cervical cancer screening [2]. A study comparing screening rates and outcomes between First Nations and all other women in Manitoba suggested lower screening rates among First Nations women aged 40 and over, and significantly higher rates of cervical lesions and cancers in First Nations women overall [34]. Targeted solutions must consider these factors. For underscreened women, the introduction of HPV self-sampling could be important in ensuring that cervical cancer screening is more accessible and acceptable [13-15, 35, 36]. A meta-analysis of 37 studies found that women who used and indicated acceptability of self-sampling for future screening did so for its convenience, privacy, and ease of use [37]. However, identified barriers included women’s lack of confidence in their ability to correctly collect the specimen and in the test result, and discomfort with the procedure [37, 38].

The concerns of *adequately screened* women (i.e., less than 3 years since previous Pap test) must also be addressed. Clear and open dialogue is needed with these women to prevent confusion and to provide reassurance that the switch to HPV-based screening is evidence-based and represents an improvement over cytology-based screening. Currently, there is a dearth of research examining Canadian women’s perceptions and understanding of HPV-based screening and potential changes to screening guidelines. To avoid problems that might arise in Canada as program implementations advance, efforts should be made to understand and address the concerns of women.

### Measuring Knowledge, Attitudes, and Beliefs About Cervical Cancer Screening

From both a theoretical and practical perspective, knowledge is a determinant of engagement in protective health behaviours [24, 39, 40]. In a systematic review of psychosocial factors associated with intentions and acceptability of HPV testing, knowledge was associated with greater acceptability of screening using the HPV test [24]. In Canada, poor cervical cancer screening knowledge has been identified as a key barrier to screening in populations such as immigrant women and ethnic minority communities [41-43]. In a recent systematic review, specific attitudes and beliefs about HPV testing have been identified as both barriers and facilitators to HPV test acceptability [24]. For example, high perceived benefits of the HPV test were associated with greater HPV test acceptability, while negative emotions (e.g., shame associated with testing for a sexually transmitted infection) related to HPV testing were associated with lower HPV test acceptability. Importantly, these findings and many additional attitudes and beliefs (e.g., high perceived susceptibility to cervical cancer, negative perceived emotional reaction to HPV test results, high perceived severity of HPV infection, etc.) have not been studied and confirmed in Canada-wide samples.

Several measures exist for cervical cancer knowledge [44-47], one subscale has been developed for measuring knowledge of HPV testing [48], and two measures of HPV testing and self-sampling attitudes and beliefs have been developed [49, 50]. However, existing measures showed insufficient psychometric testing or suboptimal psychometric properties. In addition, in a fast-moving field, many are no longer up to date with evidence from the literature on cervical cancer and HPV testing or do not include other important factors associated with HPV test acceptability (e.g., negative emotions related to HPV testing [24]). As women’s perceptions of cervical cancer and HPV testing are multifaceted, comprehensive and valid measurement tools are crucial to identifying attitudes, beliefs, and knowledge gaps that could be barriers to acceptance and uptake of HPV-based screening.

### Measuring Preferences Regarding Cervical Cancer Screening

Preferences refer to a distinct form of attitude that give information about the relative value and ranking an individual assigns to certain options over others. Measuring preferences is important in the context of cervical cancer screening as multiple approaches (e.g., varying screening intervals, use of self-sampling) are being considered for implementation [5]. Understanding Canadian women’s preferences can provide insight into the acceptability of different screening options and illuminate any potential disconnect between women and public health regarding the implementation of HPV-based screening programs. In particular, by examining women’s preferences for screening intervals and age of screening initiation - two major points of contention observed in other program implementations [22, 51] - public health can optimize communications to address concerns and ensure acceptability of HPV-based screening guidelines [24]. Furthermore, examining the preferences of underscreened and adequately screened women separately can help inform targeted communications for these groups.

### The Current Study

The proposed study will use a multi-step approach to examine women’s knowledge, attitudes, beliefs, and preferences towards HPV testing and self-sampling. A preparatory study (Study I) will focus on the development and psychometric validation of scales measuring cervical cancer screening-related knowledge, attitudes, and beliefs. Additionally, given the challenges of investigating women’s views towards a screening approach that has not yet been implemented, this study will examine the feasibility and structure of a survey examining these factors in Canadian women. Study I will yield validated scales to administer to a larger sample of Canadian women (Study II) to estimate the associations between psychosocial factors and women’s intentions to participate in HPV-based screening programs. Furthermore, Study II will survey and compare differences in psychosocial factors and HPV test intentions among *adequately screened* and *underscreened* women; providing timely data to address emerging challenges in both groups.

### Objectives

The main objectives of *Study I (Scale and Questionnaire Development*) are as follows:

1. To develop psychometrically valid scales to assess women’s knowledge of cervical cancer and attitudes, beliefs, and knowledge about HPV testing and self-sampling.
2. To evaluate the feasibility and acceptability of using a web-based survey related to Canadian women’s knowledge, attitudes, beliefs, and preferences about new HPV testing-based cervical cancer screening programs.

The main objectives of *Study II (Expanded Population-Based Survey of Canadian Women)* are as follows:

1. To estimate differences in HPV and cervical cancer screening knowledge, attitudes, beliefs, and preferences for HPV testing between women who are *adequately screened* and those who are *underscreened*.
2. To estimate the multivariable associations between psychosocial factors (e.g., knowledge, attitudes, beliefs, socio-demographics) and intentions to use HPV testing and self-sampling in women who are *adequately screened* and those who are *underscreened*.

## Study I: Questionnaire Development and Scale Validation

### Materials and Methods

#### Theoretical Frameworks

Questionnaire development and item selection for the development of scales were guided by relevant theoretical frameworks. The selection of psychosocial factors influencing intentions to engage in HPV testing was informed by two theories: the Theory of Planned Behaviour (TPB) which suggests that attitudes and beliefs, subjective norms, and perceived behavioural control influence intentions and subsequent health behaviours [52]; and the Health Belief Model (HBM) which posits that the likelihood of behavioural change is influenced by perceived susceptibility, seriousness and threat of the disease, perceived benefits of adopting protective health behaviours (i.e., screening), cues to action (e.g., information, social influence), socio-demographics, and knowledge [53].

#### Study Design

To build robust measures, we followed a rigorous stepwise process involving a review of the scientific literature, discussions, and consensus development with our experienced team of researchers, and consultation with project collaborators and the population of interest. This process is described chronologically, separated into three steps: *Phase I*.*a* Questionnaire Development, *Phase I*.*b* Questionnaire Validation, and *Phase I*.*c* Survey Testing and Psychometric Validation. At the time of manuscript submission, *Phases I*.*a* and *I*.*b* are completed, and data analysis is ongoing for *Phase I*.*c*.

#### Phase I.a Questionnaire Development

##### Literature Search

We reviewed the literature for existing relevant scales using a validated and updated search strategy that we used for a recently published knowledge synthesis summarising factors associated with HPV test acceptability in primary screening for cervical cancer [24]. Embase, Global Health, PsycInfo, Medline, and CINAHL databases were searched from January 2017-October 2019. After removing duplicates, a total of 1,477 references were screened by title and abstract and 89 full text articles were reviewed to identify relevant scales. Our team found and reviewed thirteen scales (including four scales that were identified in past literature searches [45, 48-50, 54-62]). Our overall conclusions indicated that existing questionnaires were incomplete in the following ways: not including questions about HPV testing in cervical cancer screening (e.g., only including items related to the Pap test); not having adequate psychometric validation analyses (e.g., no factor analysis or only partial factor analysis with EFA, or no reliability testing at all); having inadequate psychometric properties (e.g., internal consistency <0.6); being unsuitable for our study design (e.g., being designed to administer verbally); and having limited sampling characteristics (e.g., only adolescents or in a specific culture or context). Nevertheless, many individual items were potentially relevant to our objectives.

##### Item Selection

To evaluate potentially relevant items together and guide their selection, a large pool of items (*n* = 781) was created, and a questionnaire structure was drafted (see Figure 1), with four potential scales: cervical cancer knowledge, HPV testing knowledge, HPV testing attitudes and beliefs, and HPV self-sampling knowledge. This large item pool included items from the reviewed scales as well as new items generated based on results of a recent mixed methods synthesis of psychosocial factors affecting HPV test acceptability [24] and a systematic review of emotional responses to testing positive for HPV [25]. Items related to sociodemographics and health behaviours were also included.

**Figure 1.**
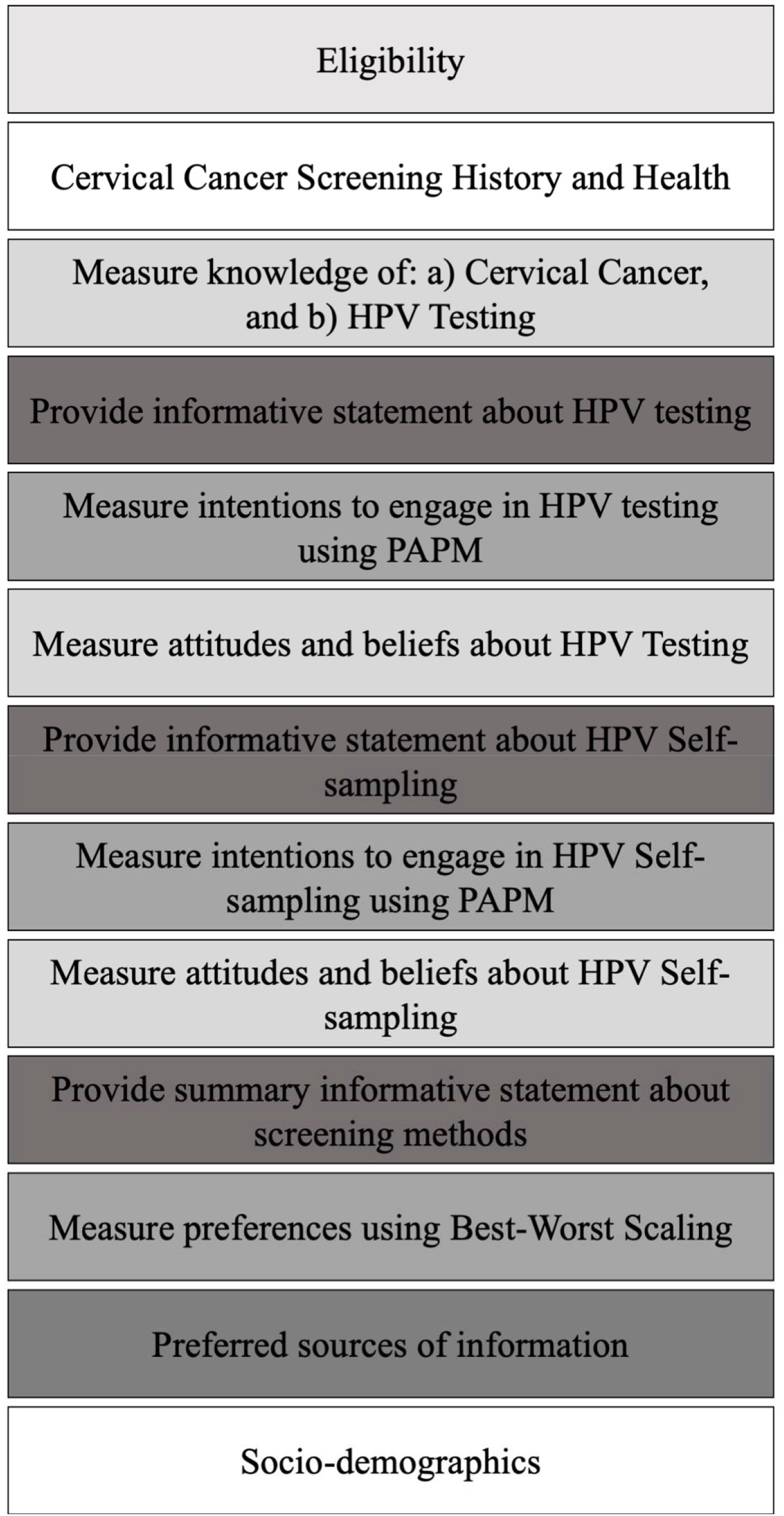
Draft Questionnaire Structure.

All 781 items were compiled into an Excel spreadsheet to be reviewed by the research team. A flow chart of item sources is found in Figure 2. Each item was reviewed individually over several research team meetings, marked as “retained” or “rejected” and categorized into an appropriate section of the questionnaire (Figure 1). For instance, the item “The HPV test is safe” was categorized as part of “Attitudes and beliefs about HPV testing” instead of “Knowledge about HPV testing” because this item captures a belief that women may agree or disagree with, and which may impact their intentions to engage in cervical cancer screening with the HPV test. Within each of the questionnaire sections, items were further mapped onto constructs from the HBM (e.g., “if the HPV test showed I have HPV, it would be serious” in *perceived seriousness*) and TPB (e.g., “my friends’ opinion about getting the HPV test would be important to me” in *social cues*) to ensure comprehensive coverage of these frameworks.

**Figure 2.**
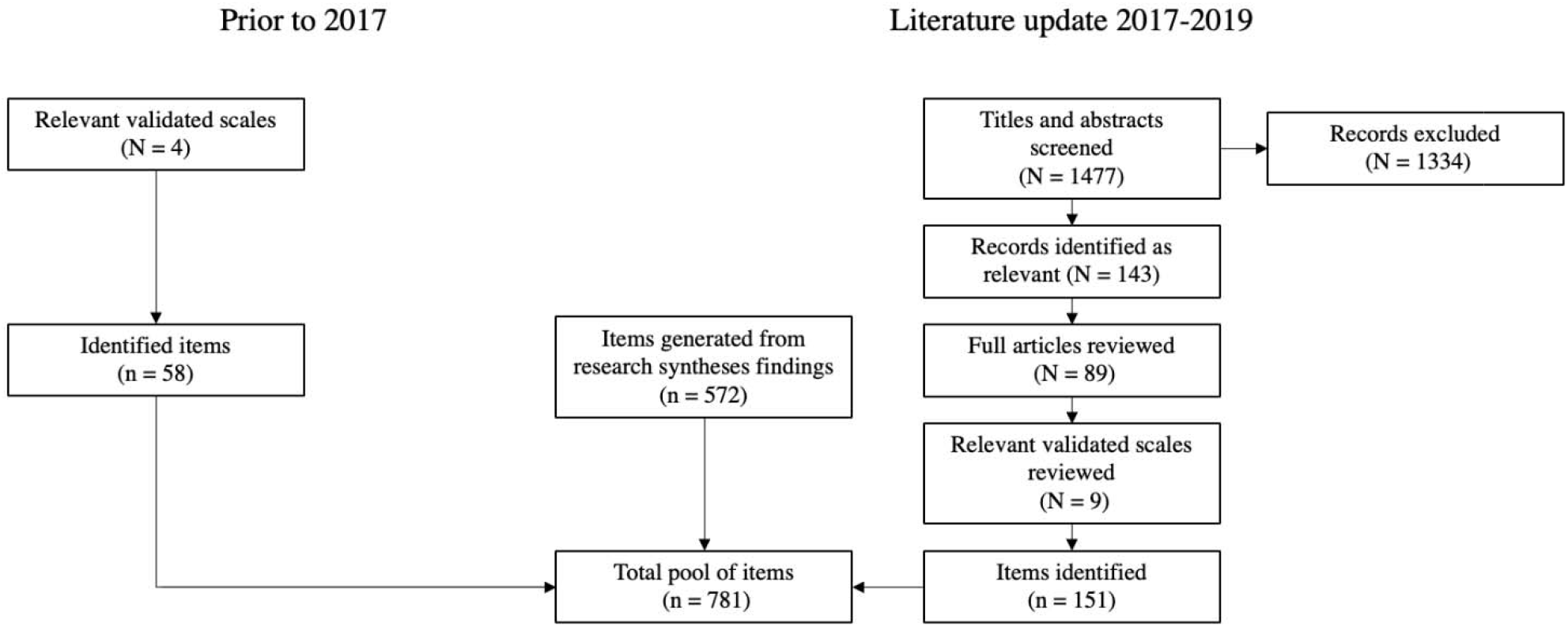
Flow Chart of Item Sources.

The focus at this stage was on the item’s underlying construct, not its wording. Reasons for rejecting items included: 1) duplicate of a factor already retained; 2) not applicable to Canadian context (e.g., “There are resources in my community for low and no cost cervical cancer screenings” as Canada has a universal healthcare system [45]); 3) infrequent, isolated factor (e.g., “I worry that tube contents may spoil/spill during transportation to doctor” [63]); and 4) item is not applicable to our project’s quantitative and survey methodology (e.g., open-ended items such as “There are many warning signs and symptoms of cervical cancer. Please name as many as you can think of:” [44]).

In total, 137 items were retained out of 781 items reviewed. Of these, 85 were categorized for scale development: 14 items for cervical cancer knowledge, 14 items for HPV testing knowledge, 44 items for attitudes and beliefs about HPV testing, and 13 items for attitudes and beliefs about HPV self-sampling. These 85 items were reviewed and revised separately for clarity, consistency, and grade eight reading level to account for different language and literacy levels. In addition, certain cervical cancer knowledge and HPV testing knowledge items were revised to achieve a balance of “True” and “False” items (i.e., made negative or affirming by adding words like ‘not’).

##### Assessing Preferences Using Best-Worst Scaling

To explore women’s preferences for their cervical cancer screening parameters (type of test, screening interval, age of screening initiation), we designed questionnaire items according to Best-Worst Scaling methodology [64]. Using this methodology, participants’ preferences were examined for different screening intervals (Domain A) or various ages of screening initiation (Domain B) while also considering multiple screening strategies (i.e., Pap test, HPV test, HPV-Pap co-testing, HPV self-sampling). In Domain A, three screening intervals were included for assessment according to their applicability to HPV test-based screening implementation: 3 years (the most common interval in Canada for existing cytology-based programs [7]), 5 years (widely recommended for HPV test-based screening [20, 65]), and 10 years (implemented for women aged 40 and over in the Netherlands and considered for wider implementation [66]). In Domain B, the following three ages of screening initiation were included: 21 years (the most common age of screening initiation in Canada for existing cytology-based programs [7]), 25 years (current recommended age of screening initiation in several Canadian provinces and widely recommended in countries implementing HPV-based screening in the context of HPV vaccination, e.g., United Kingdom and Australia [7, 20]), and 30 years (recommended age for screening initiation with primary HPV-based screening according to USPSTF guidelines [18, 67]).

##### Informative Statements

Knowledge about HPV in the general population is generally very low [68, 69], and knowledge of HPV testing and self-sampling is presumably low among Canadian women because such testing and sampling are not currently part of cervical cancer screening programs. Therefore, we sought to ensure that women had at least a basic understanding of HPV testing and HPV self-sampling before examining their attitudes, beliefs, and intentions to screen with these tests. We designed three informative statements: one about HPV testing, one about self-sampling using HPV testing, and one comparing cervical cancer screening methods. The development of these statements was inspired by brochures from other countries where HPV-based primary screening is in the process of being implemented, or already implemented (e.g., Netherlands, UK, Australia [70-72]). Each informative statement was at most one page in length, and contained a mix of text, figures, and tables (see example in **Appendix A**.). All informative statements were included *after* the items measuring participants’ knowledge.

##### Questionnaire Revisions and Translation

A draft version of the questionnaire was circulated to our team of co-investigators and collaborators (globally based researchers and public health decision-makers with expertise in HPV research, cervical cancer screening, epidemiology, oncology, psychometrics, behaviour change and health psychology theory) for a review of accuracy of content and feedback. Their suggestions were reviewed and were used to refine our questionnaire further (i.e., item wording, informative statement content, questionnaire structure). This version was then translated into French by a specialized translation firm Asiatis and verified by a native French-speaking member of the research team (GGM).

#### Phase I.b Questionnaire Validation

Given our objective of developing a comprehensible questionnaire, cognitive interviewing was used as a pre-testing method in order to detect potential sources of errors in the question-answering process [73]. This method permits an assessment of readability, understanding, and clarity of items to reveal potential differences between the intended meaning of the question by the researcher and participants’ interpretations [74]. Participants were recruited to provide verbal feedback on their understanding and experience as they completed the questionnaire over Zoom with two members of the research team present. Participants went through the items as if they were answering the questionnaire and were instructed and prompted to “think-aloud” and to provide feedback/suggestions (e.g., comment if they did not understand an item or were confused, explain how they reasoned and selected their response) [75]. Each interview lasted between 1-2 hours. Our team reviewed participants’ comments after each interview session and made changes to the questionnaire iteratively to address the issues arising.

The cognitive interviewing phase was completed when there were no new issues or feedback being raised by participants. This process required a sample of seven women, with their ages ranging between 21 and 70 years old. Four interviews were conducted in English and three were conducted in French. Based on participants’ feedback, changes were made to the questionnaire to improve the clarity of items, informative statements, instructions, page formatting, and the overall questionnaire structure and flow. See **Figure 3** for the final structure and item count.

**Figure 3.**
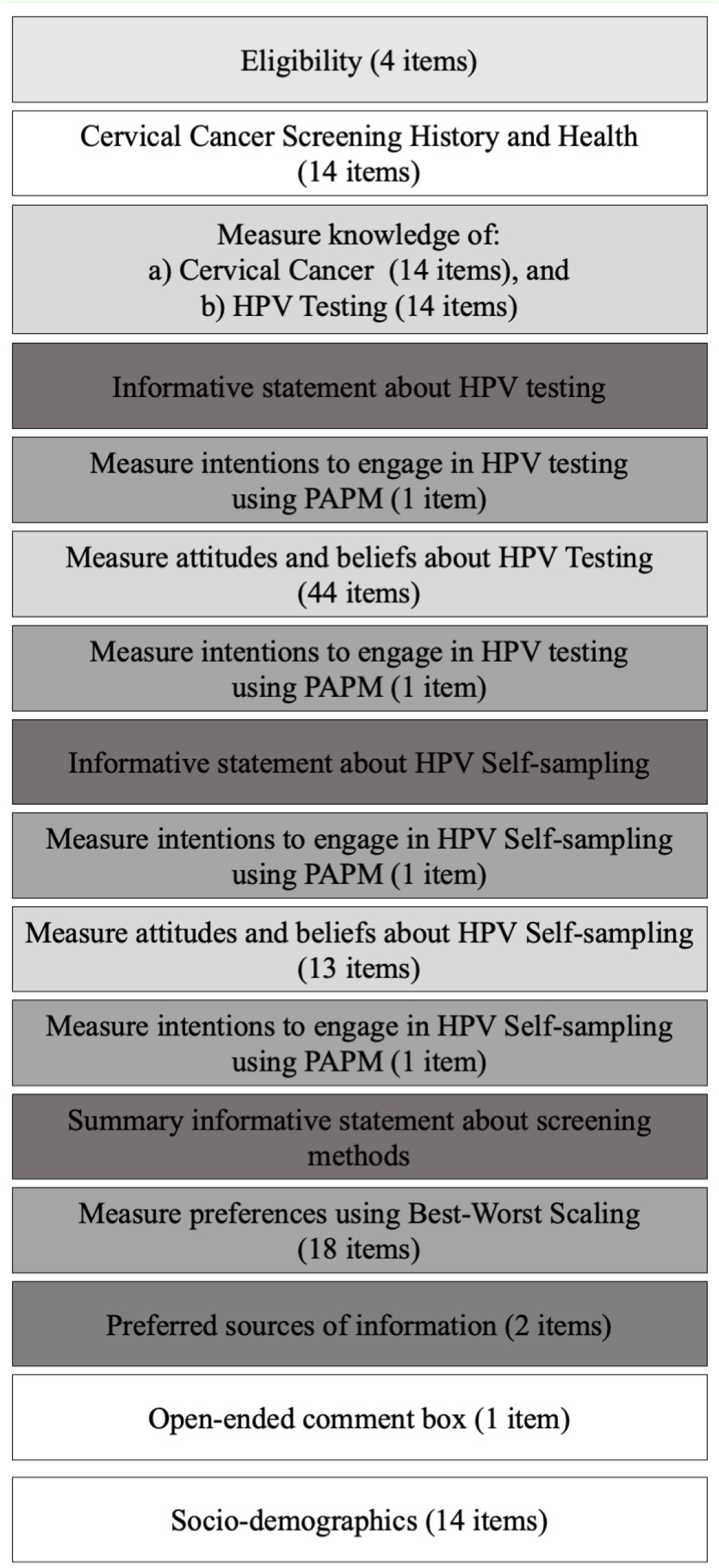
Study I Questionnaire Structure and Item Count.

#### Phase I.c Data Collection and Psychometric Validation

##### Study Design

A cross-sectional online survey was administered in October-November 2021 to a national sample of Canadian women, aged 21-70, in English and French. Participants completed the questionnaire developed in Phases I.a and I.b.

##### Participants

The inclusion criteria were as follows: 1) born female 2) between the ages of 21, the youngest age recommended to begin screening across provincial programs, and 70, the oldest age, 3) Canadian resident, and 4) intact cervix (e.g., not having undergone a hysterectomy). An exclusion criterion was having previously been diagnosed with cervical cancer. An oversampling quota was used to ensure that half of the sample were currently underscreened for cervical cancer (i.e., more than 3 years since previous Pap test, or never screened) and the other half adequately screened (i.e., less than 3 years since previous Pap test). Census-based quotas were used for primary language and province/territory of residence to reinforce sample representativeness.

##### Measures

The following measures were administered in the order presented in Figure 3.

#### Screening History

Participants were asked when they last had a Pap test to assess their screening group: a) within the last year, or b) within the last 1 to 3 years [adequately screened group], and c) over 3 years ago, or d) never [underscreened group]. Participants who reported receiving at least one previous Pap test were then asked whether they had ever received an abnormal test result. Additionally, given the study’s timing in the Fall of 2021, participants who reported receiving a Pap test over 3 years ago were asked whether the COVID-19 pandemic had prevented them from receiving a Pap test as cervical cancer screening was paused in some provinces/territories during parts of the pandemic.

#### Socio-demographics

Four items used a dichotomous response option (Yes/No) to measure identification as a visible minority [76], influence of religious or spiritual beliefs on health decisions, living in Canada for 10 years or more, and completion of a trade certificate/diploma, college degree, or university degree. Self-reported ethnic origin was measured with one item using the nine response options recommended by Statistics Canada [77]. We used multiple validated response options to measure gender identity [78]. Additionally, household income, employment status, province or territory of residence, and travel time between one’s home and a healthcare office or clinic were measured.

#### Cervical Cancer-Related Health Behaviours and Risk Factors

Participants answered the following items: self-reported health (from Very poor [1] to Excellent [6]) [79], use of oral birth control pills for 5 years or more (Yes/No), number of children given birth to (0 to 10+), smoking history (current smoker, smoked in the past, never a smoker), vaccination with at least one dose of a HPV vaccine (Yes/No/Don’t know), having a family doctor (Yes/No), height, weight, previous STI diagnosis (Yes/No), number of lifetime sexual partners, and age of sexual debut.

#### Psychosocial Items for Scale Development

Participants were presented with all items selected as part of Phase I.a. For the knowledge items, participants responded to each item with either True, False, or I don’t know. For attitudes and beliefs items, participants responded to each statement on a seven-point Likert scale from Strongly Disagree (1) to Strongly Agree (7).

#### HPV Testing and Self-Sampling Intentions

Using the Precaution Adoption Process Model (PAPM) [80], women selected their current decision-making stage with regards to the proposed screening method (HPV testing, or self-sampling using the HPV test) from five options: [1] unengaged in the decision to be screened with the HPV test/self-sampling, [2] undecided about whether to be screened with the HPV test/self-sampling, [3] decided NOT to be screened with the HPV test/self-sampling, [4] decided TO be screened with the HPV test/self-sampling, or [5] acted (already screened with the HPV test/self-sampling). A full description of the theoretical background and usage of the PAPM is provided in the “Study Design” section of Study II.

#### Best-Worst Scaling for Screening Preferences

Following the orthogonal main effects design recommended by Aizaki and Fogarty (2019) [64] and using the R software packages “DoE.base” [81] and “support.BWS2” [64], a full set of questions was developed for each of our two domains, screening intervals (Domain A) and age of screening initiation (Domain B). To examine preferences in domains with four attributes (i.e., screening methods) which each have three attribute-levels (i.e., screening interval options [Domain A], screening initiation options [Domain B]) participants must answer nine questions which contain the same number of randomly assigned combinations of attribute levels (corresponding to the defined attributes). Therefore, participants answered 18 items, nine related to cervical cancer screening intervals and nine related to age of initial screening.

#### Preferences for Screening Information and Routine Screening

Using a drag-and-drop interface, participants ranked their preferred source of information from the following options: public health agency website, social media, and a healthcare professional. A similar item was used to assess participants’ preferred type of healthcare practitioner to administer routine HPV testing amongst: family physician, gynaecologist, nurse or nurse practitioner, and physician’s assistant.

##### Recruitment and Data Collection

Recruitment was facilitated by Dynata, an international survey company with a large panel of Canadian residents recruited through a “By-Invitation-Only” method, in which participant’s identities are validated by other partnered businesses to ensure response quality. Eligible participants were invited to complete our survey through several platforms, including emails, smartphone app notifications, and Dynata’s website. Sample recruitment was conducted over two weeks in October and November of 2021. Participants were required to complete all questions before continuing the survey to prevent missing data.

##### Sample Size

We calculated the minimum sample size for conducting factor analyses using the criteria recommended by Mundform et al. (2005) that include the expected ratio of variables to factors, the level of communality, and the agreement between the sample and population solutions (K) [82]. Based on a ratio of variables to factor = 4 (for the 13 items reflecting attitudes and beliefs for sampling), a low level of communality (i.e., range of .2 to .4) and an excellent agreement between the sample and population solutions (i.e., K=0.98), the minimum sample required was 500 (rounded up from 450). Because our analytic plan includes conducting Exploratory Factor Analysis (EFA) and Confirmatory Factor Analysis (CFA) on separate samples and considering that oversampling is needed to account for approximately 18% invalid and inattentive responses [83] the sample needed is 1220 observations (i.e., 2*500*100/82=1219).

##### Data Cleaning

Direct and statistical data cleaning methods were used stepwise to identify potentially “inattentive” or “unmotivated” respondents in our final dataset. Two attention check items were used in the survey, one within the items related to HPV test attitudes and beliefs scale, and one in the items related to HPV self-sampling attitudes and beliefs. Following the instructed response design [83], each of these items asked participants to select a specific response choice from a Likert scale to verify attention (e.g., “please select “strongly agree”, for this question only”). Participants who responded correctly to at least one of these items were retained. Next, participants who “straight-lined” (i.e., answered all items using the same response) were identified by calculating the variance in response for all the HPV testing attitudes and beliefs items. Participants who exhibited no variance in their responses across these items were excluded. Finally, those in the longest 2.5% and shortest 2.5% of survey response times in the remaining participants were excluded. In total, N=1027 valid responses were retained for analyses.

### Data analysis

#### Objective 1. Scale Validation

To evaluate dimensionality, we will conduct Exploratory Factor Analysis (EFA) and Confirmatory Factor Analysis (CFA) separately for cervical cancer knowledge, HPV testing knowledge, and HPV testing and self-sampling attitudes and beliefs items. The final dataset will be split randomly into two equal samples and one sample will be used to conduct EFA and the other to validate the factor structure using CFA. To select the optimal number of factors we will use the parallel analysis approach and the syntax developed by O’Connor [84]. For items within each factor, we will use Item Response Theory modelling; for binary data (knowledge items) we will use the two-parameter logistic regression model that accounts for item difficulty and discriminant capacity, and for ordinal data (attitudes and beliefs items) we will use the graded response model that accounts for discriminant capacity and probability of selecting a certain Likert scale score (e.g., *strongly agree*). Concurrently, we will examine how well each item measures the latent trait by plotting item information against the latent construct ability. We aim to retain items that cover a wide range of difficulty and that have high discriminant capacity and information value. To estimate the CFA model fit we will use the fit indices and (cut-off criteria) recommended by Hooper et al. (2008) [85]: (a) Wheaton et al.’s relative/normed chi-square (χ2/df =2 to 5), (b) the standardized root mean square residual (SRMR; <.8), (c) the root mean square error approximation (RMSEA;<.06), (d) the comparative fit index (CFI; ≥.95), and (e) the Tucker-Lewis index (TLI; ≥.95). To evaluate internal consistency for items loading onto each factor, Cronbach’s α will be calculated.

#### Best-Worst Scaling Preferences

To analyze BWS preference data, we will use the counting and the modelling approaches described by Aizaki and Fogarty (2019) [64]. In line with the counting approach, for domain A (screening interval) and domain B (age of initiation) we will calculate the best-minus-worst (BWs) score (higher scores reflect higher preferences) for each attribute (e.g., HPV test) and attribute level [e.g., three years (for screening interval); 25 years (for age of initiation)]. For the modelling approach, we will use conditional logistic regression to model preferences as a function of the sum of the value of attributes and attribute-levels [64]. For attributes and attribute levels we will estimate odds ratios and 95% confidence intervals.

### Objective 2. Questionnaire Feasibility

Preliminary results suggest a high level of feasibility. Through Dynata’s recruitment methods, data collection was completed within two weeks. As detailed in “Data Cleaning”, overall, data was of good quality, with the exclusion of 16.5% of complete responses for poor quality being consistent with previous research in our lab and expectations for survey research [39, 83, 86]. The mean response time (post data cleaning) was 30.4 minutes, consistent with other surveys conducted by our research team [87]. It is possible that some participants left the survey and returned to it later, which could explain the slightly higher response time than our estimated 25 minutes. Given that scales in the Study II will be made more concise through psychometric analyses applied to responses from this study, we expect this survey will be shorter in length. Of a total of 1392 participants who were eligible and began the survey, 1242 completed the survey, an attrition rate of 10.77% (see Appendix B. for a diagram detailing participant attrition). Attrition primarily occurred in the HPV attitudes and beliefs (n = 50), the longest section of the survey with 44 items. We expect that the validation and shortening of this scale for Study II will help to address this issue.

#### Study I Ethics

Study I received ethical approval from the Research Ethics Board of The Integrated Health and Social Services University Network (CIUSSS) West-Central Montreal (Project ID: 2021-2632).

## Study II: Expanded Population-Based Survey of Canadian Women

### Materials and Methods

#### Study Design

A cross-sectional online survey will be administered in Spring-Summer 2022 to a nationally representative sample of Canadian women, aged 21-70, in English and French. The study population and inclusion criteria will be the same as that recruited in Study I. Oversampling will be used to ensure half of the sample is adequately screened, and the other half is underscreened. Additional quotas will be used for the following factors: age, household income, and rural/urban residence (considering lower cervical screening rates in rural Canada [7]) based on Statistics Canada census data. Data collection will be conducted by Dynata following the same procedure as in Study I.

The questionnaire will follow the same structure and include the same sections as the survey detailed in Study I. Of the knowledge, attitudes, and beliefs items used for scale development in Study I, only those items retained after extensive psychometric analyses will be used in the Study II questionnaire as part of the resulting shorter, validated scales. Two previously validated scales, the extended HPV General Knowledge and HPV Vaccine Knowledge, will be added [48, 69], given the relevance of HPV and HPV vaccination to cervical cancer. The measurement of the outcome variables (intentions to screen using the HPV test and self-sampling) is informed by the Precaution Adoption Process Model (PAPM) [80], a categorical, stage-based model of health decision-making. Whereas a binary, “Yes/No” outcome limits accuracy, using a multi-stage model provides a more precise, nuanced understanding of women’s decision-making process that acknowledges the unique barriers associated with movement between each stage towards engaging with the behaviour [88].

#### Sample Size

We calculated the sample size based on the estimation that approximately 30% of women will be in each of the PAPM adoption stages *unengaged, undecided*, and *decided to* and 10% in the stage *decided not*. Because multinomial logistic regression can be considered a series of binary logistic regressions, we based our calculations on the work of Peduzzi et al. (1995) [89], who recommends a minimum of 10 observations per variable and the formula N = 10k/p where N = minimum number of observations needed, k = number of predictor variables and p = smallest of the proportion in the binary model (i.e., N = 10*15/0.25 = 600). Therefore, the minimum sample needed to reach enough power is 1500 because 40% of the sample (e.g., unengaged + decided NOT) must represent 600 observations. Given that our objective is to estimate the association between psychosocial factors and intentions of HPV testing in both underscreened and adequately screened women, we need 3000 (i.e., 1500 in each group) valid responses to perform the analyses. Considering that an oversampling of maximum 18% is required to account for careless responses, the approximate total number of survey responses needed is 3650.

#### Data Analysis

Corresponding to Objective 1, we will conduct univariate analyses (means and proportions) for scale items (e.g., knowledge, attitudes) and bivariate analyses (t-tests and Chi-square) to estimate differences between adequately screened and underscreened women. We will calculate the effect size using Cohen’s h and Cohen’s d for proportions and continuous variables respectively. Corresponding to Objective 2, we will estimate the associations between psychosocial factors and intentions of HPV testing and self-sampling using multinomial logistic regression and model the log odds of PAPM stages (dependent variable) as a linear combination of the independent variables (e.g., HPV testing knowledge, BWS preference scores). We will report odds ratios (OR) and 95% confidence intervals of being in a PAPM stage versus the reference category (i.e., *unengaged*) for each independent variable. Initially, we will conduct bivariate analyses to estimate the association between the outcome and independent variables. Then, independent variables showing significant associations (p<0.1) will be entered simultaneously in the final model. We selected the following indices to report model fit: (a) Cox-Snell R2, (b) Cragg-Uhler R2 and (c) McFadden R^2^ [39, 90]. We will use the Hausman-McFadden test to evaluate the final model for independence of irrelevant alternatives (IIA) which postulates that a person’s choice (i.e., PAPM stage) is unchanged by other available choices (i.e., fewer PAPM stages) [91]. Analyses will be conducted separately for underscreened and adequately screened women. Analyses will be conducted in SPSS and STATA.

#### Study II Ethics

Study II has received ethical approval from the Research Ethics Board of the CIUSSS West-Central Montreal (Project ID: 2022-2960).

## Discussion

### Study Implications

Understanding the psychosocial factors that might affect women’s perceptions of and intentions to pursue screening with HPV-based screening will be critical as Canada plans to implement changes to cervical screening programs and guidelines. Through this multi-step study, we will develop several validated scales to facilitate population-based investigations of these factors by other researchers. The use of these scales to investigate a representative sample of Canadian women’s perceptions of HPV-based screening will provide directly applicable knowledge to public health and healthcare professionals.

Scales developed in Study I will be informed by measures in the extant literature, and further enhanced by the generation of new items based on systematic evaluation of themes in the psychosocial literature on HPV testing. Using the TPB and HBM to guide item selection will further ensure scales provide meaningful insights into factors impacting screening behaviours. As a result, the developed measures will provide a comprehensive assessment of attitudes, beliefs, and knowledge relevant to cervical cancer screening, and in particular HPV test-based screening. Application of advanced psychometric methods will enable the evaluation and optimization of our scales to accurately examine the constructs in question. We expect our scales to act as a new measurement standard in investigating attitudes, beliefs, and knowledge associated with HPV-based screening. Additionally, use of Best-Worst Scaling to estimate preferences for cervical cancer screening will enable an evaluation of women’s perceptions of trade-offs between different testing methods, and importantly, screening intervals and age of initial screening. These changes have been barriers in other countries where HPV-based screening has been implemented [22, 92].

Our expanded population-based survey (Study II) will provide comprehensive data to inform and support the development of Canadian HPV-based cervical cancer screening programs. Through objective 1, the specific evaluation of differences in knowledge, attitudes, and beliefs about HPV testing among women who are underscreened or adequately screened will enable the development of targeted interventions to improve knowledge, address negative attitudes and beliefs, and reassure women about upcoming changes. In objective 2, analyzing the sociodemographic and psychosocial factors associated with screening intention stages will provide insight into developing interventions that consider the multiplicity of barriers to screening. Our investigation will offer a valuable response to calls to action to examine inequalities in cervical cancer screening in the interest of cervical cancer elimination [8]. Further, by comparing women’s knowledge, attitudes, beliefs, and preferences towards HPV-based screening with proposed guidelines, our findings will help to inform screening recommendations and ensure a successful transition from cytology to HPV-based screening in Canada.

### Knowledge Dissemination Plan

To reach both research and health professional audiences, study findings will be published in open access, peer-reviewed scientific journals. Presentations will be made at national and international scientific meetings (e.g., Canadian Association of Psychosocial Oncology, International Papillomavirus Society (IPVS) conference, the International Psycho-Oncology Society) and in online webinars (e.g., Consortium for Infectious Disease Control; IPVS). To reach policymakers, we will share a final research report with the Public Health Agency of Canada (PHAC), Canadian Partnership Against Cancer (CPAC), Canadian Task Force on Preventive Health Care (CTFPHC), and provincial/territorial cancer screening program advisory boards. We will also share our results with nonprofit organizations such as the Canadian Cancer Society, the Society of Obstetricians and Gynaecologists of Canada (SOGC), the College of Family Physicians of Canada, and HPV Global Action who have expressed strong interest in disseminating our results. We will produce content (e.g., infographics) and engage with Canadian women on social media (Twitter, Facebook, etc.). Research summaries will be drafted for dissemination to national media outlets to inform women about this public policy change and encourage discussions about HPV-based cervical cancer screening.

### Study Strengths

A major strength of this study is the rigorous and comprehensive process to develop psychometrically validated scales informed by theoretical frameworks. Additionally, examination of the feasibility of the developed questionnaire through Study I will ensure that the survey used in Study II will be comprehensible and relevant to Canadian women. Best-Worst Scaling presents an innovative approach to assess preferences that controls for biases observed in typical multiple-choice or ranking assessments of preferences. The use of the PAPM to examine intentions towards HPV testing and self-sampling will provide a theoretically informed and nuanced understanding of women’s decision-making compared to other studies using continuous or binary measures. Using a market research polling firm will enable time- and cost-efficient recruitment and data collection. Additionally, online survey methodology will prevent the issue of missing data. Quota-based sampling will allow us to recruit a nationally representative sample using based on recent census data. Attention check items and data cleaning techniques will allow us to identify and exclude unmotivated or “careless” responders, ensuring that high-quality data will be collected and analyzed.

### Foreseeable Limitations

Relying on respondents’ self-report for their screening history of having had a Pap test in the last three years or not is subject to recall bias. Our anonymous online survey design prevents us from verifying it against health records data. To minimize this limitation, women were provided with a reminder informative statement explaining what a Pap test is and how it is performed before asking them about their screening history. Given that the COVID-19 pandemic has prevented and continues to prevent women from engaging in cervical cancer screening, this may affect women’s report of their screening history and change the composition of women in our “underscreened” and “adequately screened” categories. It is not clear how this will affect our data as the COVID-19 pandemic’s effect on screening access in Canada is not well understood in most provinces and territories [93]. To address this issue, we will include an item asking those participants who report being underscreened whether the COVID-19 pandemic had prevented them from receiving screening, and sensitivity analyses will be performed to examine the pandemic’s impact on screening. Additionally, the lack of an investigation at two different time-points precludes any investigation of how intentions for HPV testing and self-sampling might relate to uptake and acceptability as these testing methods are introduced. Longitudinal examinations of acceptability and intentions towards HPV-based screening will be needed as implementation occurs in Canada. Unfortunately, our study design and recruitment strategy might preclude specific analyses in gender and sexual minority groups, considering low case counts observed in population-wide web-surveys (e.g., 0.4% gender diverse in Study I). Recognizing the unique barriers faced by these groups in screening for cervical cancer, future research should specifically investigate the perceptions of gender and sexual minority Canadians towards HPV testing and self-sampling implementation [3, 14, 94]. Similarly, a comprehensive understanding of cervical cancer screening and HPV in First Nations, Inuit, and Métis populations is critical [5, 34]. Our study could highlight certain issues in these populations (considering 2.9% representation in Study I), but unique participatory action initiatives involving relevant NGOs and community advocacy groups are needed to fully understand their views and address their concerns.

## Data Availability

All data produced in the present study are available upon reasonable request to the authors and depending on the guidelines set out by the approving research ethics board.

## Acknowledgements

The authors thank Eduardo Franco (McGill University) for his feedback on the project aims, design, and survey content. We also thank Johnny Dagher (Dynata) and his colleagues for their expert help in programming and administering the survey for this study.

## Appendix A. Example Informative Statement

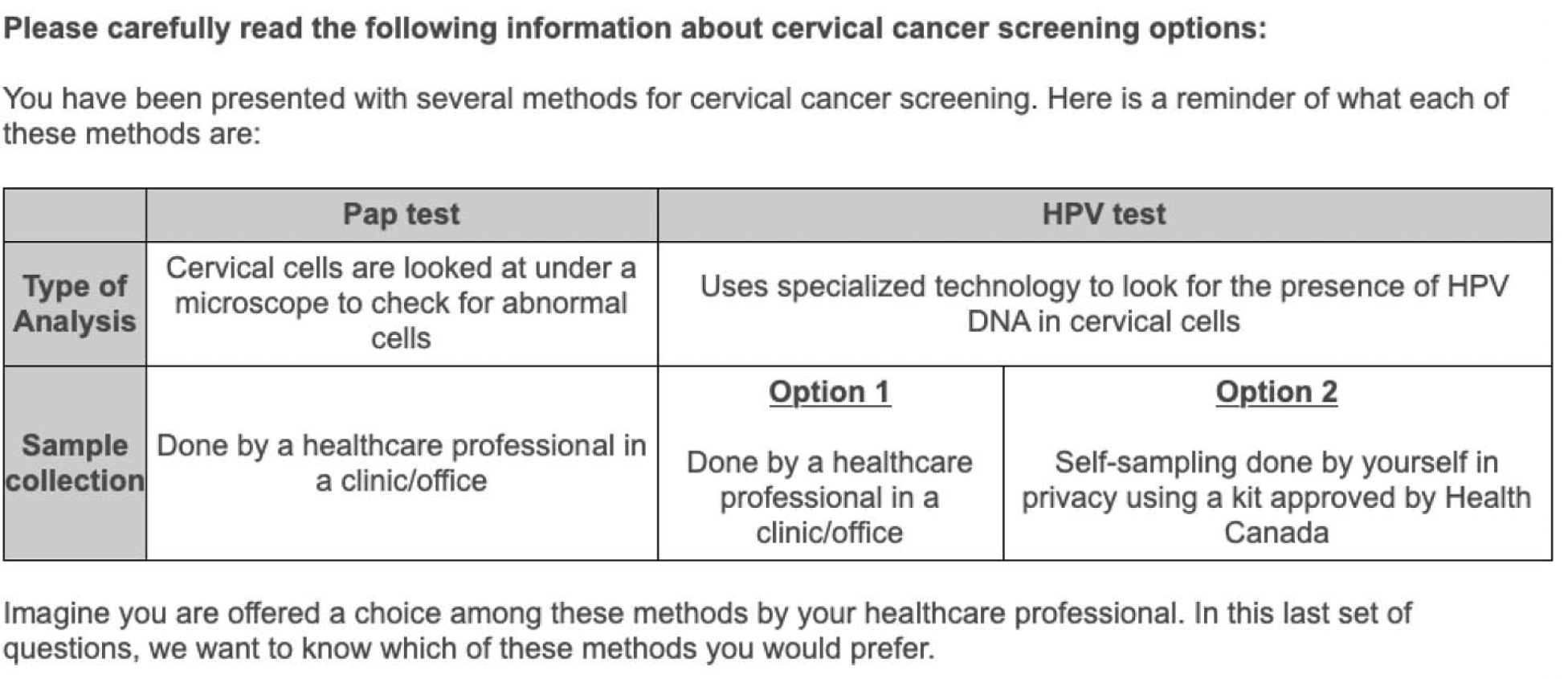

## Appendix B. Study I Participant Flow Diagram

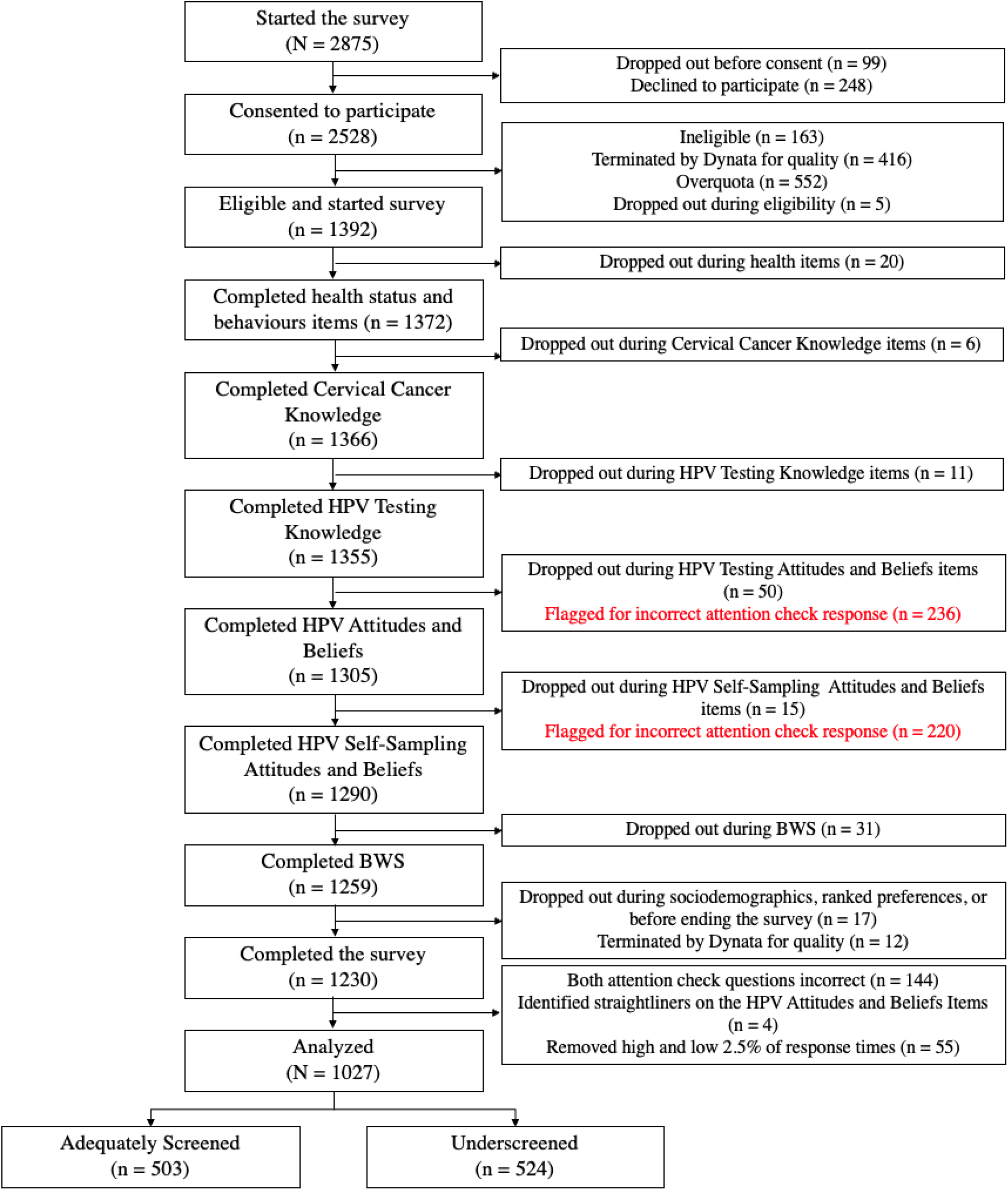

